# Snoring and risk of dementia: a prospective cohort and Mendelian randomization study

**DOI:** 10.1101/2023.10.12.23296972

**Authors:** Yaqing Gao, Shea Andrews, Willa Brenowitz, Cyrus A Raji, Kristine Yaffe, Yue Leng

**Author notes:** Joint first authors (Yaqing Gao, Shea Andrews). Correspondence to: Yue Leng, Department of Psychiatry and Behavioral Sciences, University of California San Francisco San Francisco, CA 94107, USA.

## Abstract

**Background:** The association between snoring, a very common condition that increases with age, and dementia risk is controversial. Snoring is linked to obstructive sleep apnoea and cardiometabolic conditions, both of which are associated with an increased risk of dementia. However, snoring also increases with body mass index (BMI), which in late life is linked to lower dementia risk, possibly due to metabolic changes during prodromal dementia.

**Methods:** The prospective cohort study used data from 450,027 UK Biobank participants with snoring measured at baseline (2006 – 2010), and followed up for dementia diagnosis (censored at 2022). Two-sample Mendelian randomization (MR) analysis used summary statistics for genome-wide association studies of Alzheimer’s disease (AD) (n = 94,437; cases = 35,274) and snoring (n = 408,317; snorers = 151,011).

**Results:** During a median follow-up of 13.5 years, 7,937 individuals developed dementia. Snoring was associated with an 8% lower risk of all-cause dementia (hazard ratio [HR] 0.92; 95% confidence interval [CI] 0.88 to 0.97) and AD (HR 0.92; 95% CI 0.86 to 0.99). The association was stronger in older individuals, *APOE* ε4 allele carriers, and during shorter follow-up periods. MR analyses suggested no causal effect of snoring on AD, however, genetic liability to AD was associated with a lower risk of snoring. Multivariable MR indicated that the effect of AD on snoring was primarily driven by BMI.

**Conclusions:** The phenotypic association between snoring and lower dementia risk likely stems from reverse causation, with genetic predisposition to AD associated with reduced snoring. This may be driven by weight loss in prodromal AD.

## Key Messages

**What was your research question?**

Is there a causal association between snoring and dementia risk, and, if so, does body mass index play a role?

**What did you find?**

We found that snoring was associated with a lower risk of incident all-cause dementia and Alzheimer’s disease (AD). These associations were stronger in older adults and *APOE* ε4 carriers and were attenuated with increasing follow-up length, indicating potential reverse causality. Mendelian randomization analysis confirmed a causal association between AD and reduced snoring, which was driven by decreased body mass index as an AD prodrome.

**Why is it important?**

Increased attention should be paid to reduced snoring and weight loss in older adults as potential early indicators of dementia risk.

## Introduction

Snoring, a very common condition affecting 35-45% of males and 15-28% of females in the general population,^1^ is a noise that arises from increased resistance to airflow in the upper airway during sleep and vibrations of the surrounding tissues.^2^ Snoring increases with age and body mass index (BMI), and is frequently associated with obstructive sleep apnoea (OSA) and cardiometabolic diseases.^3–5^ However, the research evidence regarding the association between snoring and dementia is scarce and controversial.

While a case-control study suggested that demented patients snored twice as frequently as control subjects,^6^ two meta-analyses found no association between snoring and risk of dementia,^7, 8^ and one study suggested higher cognitive function associated with snoring.^9^ There are several explanations for these inconsistent findings. First, a positive association, possibly bi-directional, may exist. Patients with dementia experience exacerbated age-related neuromuscular changes, such as heightened airway collapsibility or reduced airway muscle responsiveness, increasing the likelihood of snoring.^10^ Snoring may be associated with OSA and cardiometabolic diseases, which have been suggested as risk factors for dementia.^11–13^ Second, snoring is associated with vascular damage, possibly through hypoxia-related inflammation and oxidative stress,^14^ as well as pressure waves from snoring vibrations transmitted to the carotid arterial wall.^15^ This may contribute to the risk of vascular dementia (VaD) rather than Alzheimer’s disease (AD). Therefore, failure to consider the subtypes of dementia or focusing on AD as the outcome may lead to null findings. Third, while obesity is one of the strongest predictors for snoring,^1^ low BMI in late life has been associated with an increased risk of AD,^16^ which could be due to metabolic alterations and decreased intake in prodromal AD.^17^ This may lead to a negative association between snoring and risk of AD, especially in older individuals. Therefore, it is crucial to unravel the causal relationship between snoring and dementia, the direction of the relationship, and the role of BMI in this link.

Mendelian randomization (MR) employs single-nucleotide polymorphisms (SNPs) as genetic instrumental variables for the proposed risk factor that affects health. Therefore, it can be used to address some of the limitations of observational studies, particularly confounding and reverse causation. Multivariable MR (MVMR) estimates the direct effect of the genetic liability of the exposure on the outcome, while adjusting for the genetic liability of a second exposure of interest.^18^ MR and MVMR have yet to be applied to snoring, BMI, and dementia, which could help disentangle the causal relationship among these factors.

In this study, we aimed to determine the association of snoring with incident all-cause dementia, AD, and VaD, using a conventional observational study approach in a large longitudinal cohort of over 500,000 middle-to older-aged adults, and a two-sample bi-directional MR design to elucidate the causal relationship between snoring and AD. We also utilized MVMR analyses to examine the role of BMI in the link between snoring and AD.

## Methods

### Study population

This prospective cohort study used data from UK Biobank, which recruited over 500,000 individuals aged 40 to 69 years from 22 assessment centres across the UK between 2006 and 2010.^19^ Baseline assessments included touch-screen questionnaires, verbal interviews, physical measurements, and genotyping blood samples. Follow-up information was obtained through linkage to hospital inpatient records and death registries from national datasets in England, Scotland, and Wales.

For the current study, out of the 502,406 participants initially recruited, we excluded those who self-reported dementia at baseline or had a hospital inpatient record of dementia prior to baseline (n=230). Additionally, 52,186 participants with missing exposure or covariate data were excluded from the main analysis but included through multiple imputation in sensitivity analysis. The final sample size for the main analysis consisted of 450,027 participants.

### Assessment of snoring

At baseline, participants provided information on sleep-related traits. Snoring was assessed using a single item (Field-ID: 1210): “Does your partner or a close relative or friend complain about your snoring?” Participants were given the following response options: “Yes,” “No,” “Don’t know,” or “Prefer not to answer.” Participants who selected “Don’t know” or “Prefer not to answer” were categorized as having missing data.

### Diagnoses of Incident Dementia

Dementia diagnoses were determined by using the UK Biobank-linked hospital inpatient records and death registries, which were documented based on the International Classification of Disease (ICD) codes (e-Table 1). Individuals who received a primary or secondary diagnosis of dementia after the baseline assessment or had dementia identified as an underlying or contributory cause of death were categorized as having incident dementia. The primary outcome of the present study was all-cause dementia, while subtypes of dementia, including AD and VaD, were assessed as secondary outcomes.

### Statistical Analysis

#### Prospective Cohort Study Analysis

We first utilized data from the UK Biobank to assess the longitudinal association between snoring and the subsequent risk of dementia. Participants were followed from baseline until the date of first dementia diagnosis, death, loss to follow-up, or the censor dates for hospital inpatient data (October 31, 2022 for England; July 31, 2021 for Scotland; and February 28, 2018 for Wales), whichever occurred first. We applied Cox proportional-hazards models with follow-up time as the underlying time scale, and checked the proportional hazards assumption through the Schoenfeld residual tests. Models were adjusted sequentially for age, sex, ethnicity, education, and TDI quintiles (Model 1); smoking status, alcohol consumption, daytime dozing, BMI, histories of depression, diabetes, hypertension, and cardiovascular diseases, which were obtained through baseline verbal interviews and hospital diagnoses prior to baseline (Model 2). The secondary outcomes (i.e., AD and VaD) were analysed using the same models. Detailed definitions and classifications for covariates and ICD codes used in detecting disease diagnoses are listed in e-Table 1-2.

To examine potential effect modification by factors such as age, sex, and genetic susceptibility to dementia (i.e., *APOE* ε4), we incorporated interaction terms between snoring and each of these modifiers and conducted stratified analyses.

To examine the robustness of our findings, we performed two sensitivity analyses. Firstly, the analysis was stratified based on the duration of follow-up, including participants who were followed for ≤5 years, 5 to 10 years, and >10 years after the baseline assessment. Secondly, participants with a history of sleep apnoea at baseline were excluded from the analysis. Thirdly, we performed multiple imputation to impute missing exposure and covariate data (e-Appendix 1).

#### MR Analysis

We utilized the latest GWAS summary statistics for snoring, which involved over 400,000 individuals from the UK Biobank, including more than 150,000 snorers, and identified 42 loci that were genome-wide significant.^1^ We obtained GWAS summary statistics for Alzheimer’s disease (AD) from the study based on International Genomics Alzheimer’s Project, which reported 20 genome-wide significant loci in a sample of 94,437 individuals.^20^ GWAS summary statistics for BMI were obtained from the most recent GWAS meta-analysis, which included association findings for up to 339,224 individuals and had no overlapping sample with the AD GWAS, identifying 97 loci.^21^ To identify instruments for each exposure, independent genome-wide significant SNPs were extracted (pLJ<LJ5LJ×LJ10^−8^) from their respective GWASs. Details for clumping, proxy identification, and harmonization of effect sizes for the instruments on outcomes and exposures can be found in eMethods. The harmonized datasets are available in e-Table 3.

The primary MR method employed to assess causal association was the fixed-effects inverse variance weighted (IVW) method, which assumes all genetic variants are valid instruments – that is, they don’t violate any of the underlying assumptions for MR. We conducted sensitivity analyses using alternative methods known to produce more robust causal estimates in the presence of horizontal pleiotropy but at the cost of reduced statistical power. These methods included MR-Egger regression, Weighted Median Estimator (WME), and Weighted Mode Based Estimator (WMBE). We conducted a multivariable Mendelian Randomization (MVMR) analysis to assess whether the causal association remains significant when controlling for the effect of BMI. Diagnostics tests included the MR-Egger regression to evaluate the presence of directional horizontal pleiotropy, Cochran’s Q test to evaluate heterogeneity, Radial MR to identify outliers, and F-statistics to evaluate instrument strength. Figure 1 presents the hypothesized pathways examined through univariate and multivariate MR.

**Figure 1.**
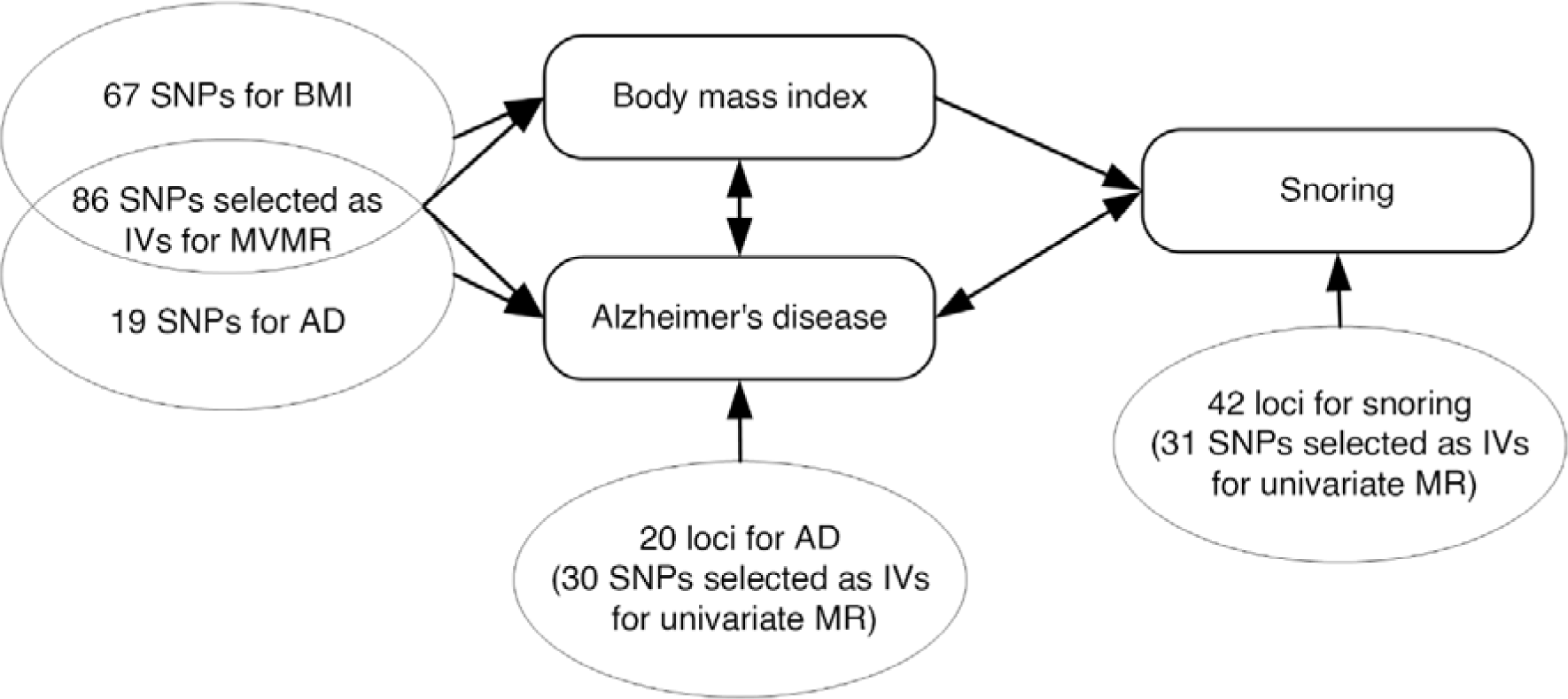
Hypothesized pathways examined through univariate and multivariable Mendelian randomization. MR, Mendelian randomization; MVMR, multivariable Mendelian randomization; SNPs, single nucleotide polymorphisms; BMI, body mass index; IVs, instrumental variables; AD, Alzheimer’s disease.

## Results

### Prospective Cohort Analysis

Among the 450,027 participants, 167,662 (37.3%) reported snoring (Table 1). Compared with non-snorers, snorers tended to be older, more often male, and current smokers, and had higher alcohol consumption. In addition, snorers had a higher BMI and were more likely to have a history of daytime dozing, depression, hypertension, cardiovascular diseases, and diabetes.

**Table 1.** Baseline characteristics of participants by snoring status.

| **Characteristic** | **Overall** | **Not snoring** | **Snoring** |
| --- | --- | --- | --- |
| **N** | 450027 | 282365 | 167662 |
| **Age (mean (SD))** | 56.97 (8.10) | 56.80 (8.28) | 57.25 (7.78) |
| **Female (%)** | 242527 (53.9) | 173861 (61.6) | 68666 (41.0) |
| **Townsend deprivation index quintile** |  |  |  |
| 1 (Least deprived) | 93392 (20.8) | 58126 (20.6) | 35266 (21.0) |
| 2 | 92326 (20.5) | 57378 (20.3) | 34948 (20.8) |
| 3 | 90984 (20.2) | 56933 (20.2) | 34051 (20.3) |
| 4 | 89063 (19.8) | 56229 (19.9) | 32834 (19.6) |
| 5 (Most deprived) | 84262 (18.7) | 53699 (19.0) | 30563 (18.2) |
| **Education (%)** |  |  |  |
| Primary | 75849 (16.9) | 46459 (16.5) | 29390 (17.5) |
| Secondary | 101074 (22.5) | 64131 (22.7) | 36943 (22.0) |
| Post-secondary non-tertiary | 54427 (12.1) | 34190 (12.1) | 20237 (12.1) |
| Tertiary | 218677 (48.6) | 137585 (48.7) | 81092 (48.4) |
| **Non-white (%)** | 22054 (4.9) | 13713 (4.9) | 8341 (5.0) |
| **Drinking daily or almost daily (%)** | 93586 (20.8) | 55109 (19.5) | 38477 (22.9) |
| **Current smoker (%)** | 46042 (10.2) | 26371 (9.3) | 19671 (11.7) |
| **Obese (%)** | 108341 (24.1) | 53548 (19.0) | 54793 (32.7) |
| **Daytime dozing (%)** | 106616 (23.7) | 59189 (21.0) | 47427 (28.3) |
| **Medical history (%)** |  |  |  |
| Depression | 26522 (5.9) | 15836 (5.6) | 10686 (6.4) |
| Hypertension | 121889 (27.1) | 68023 (24.1) | 53866 (32.1) |
| Cardiovascular diseases | 44068 (9.8) | 25924 (9.2) | 18144 (10.8) |
| Diabetes | 22336 (5.0) | 12026 (4.3) | 10310 (6.1) |
| **APOE ε4 carrier (%)** | 114256 (26.6) | 72050 (26.8) | 42206 (26.3) |

N, number of participants; SD, Standard deviation; BMI, body mass index.

During a median follow-up of 13.5 years, we identified 7,937 incident cases of dementia. Among these cases, 3,515 were diagnosed as AD, and 1,710 as VaD. In Cox regression analysis adjusted for sex, age, and socioeconomic status, snoring was associated with a lower risk of incident all-cause dementia (hazard ratio [HR], 0.94; 95% confidence interval [CI], 0.90 to 0.98) (Figure 2). Secondary analysis of dementia subtypes revealed that snoring was associated with a lower risk of AD (0.91, 95% CI 0.85 to 0.98), whereas no significant association was found for VaD. The magnitude of association between snoring and dementia remained largely unchanged after adjusting for the lifestyle factors and health status. In the fully adjusted model, snorers had an 8% lower risk of developing all-cause dementia and AD compared to non-snorers (0.92, 95% CI 0.88 to 0.97 and 0.92, 95% CI 0.86 to 0.99).

**Figure 2.**
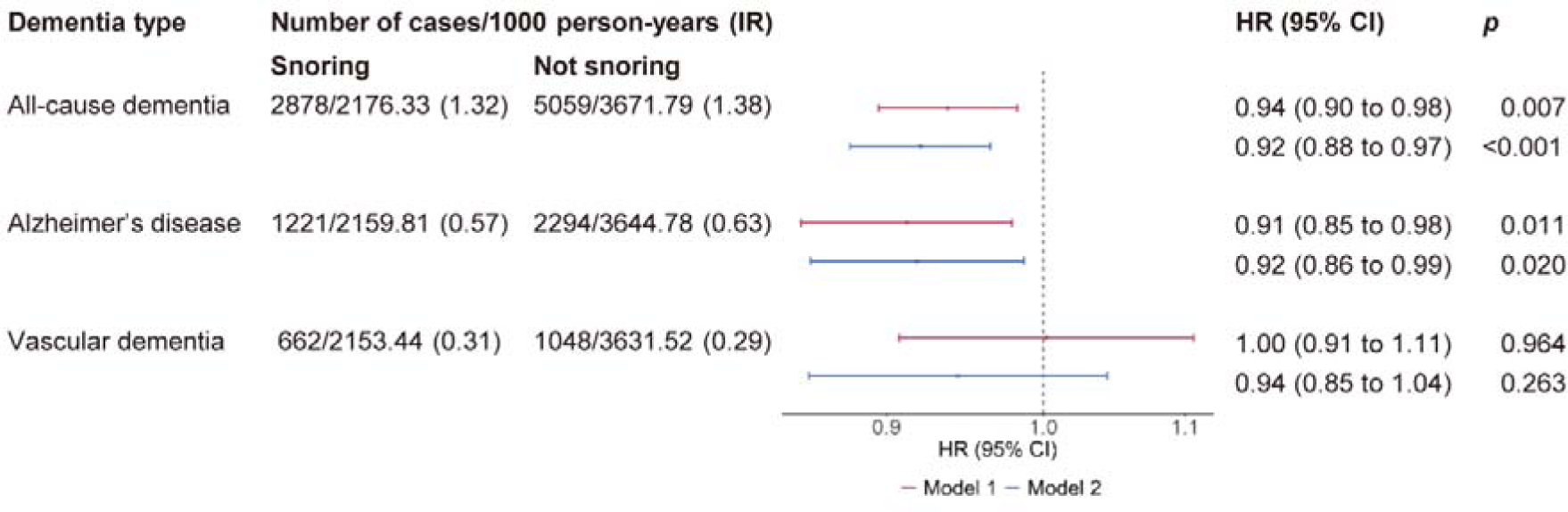
Multivariable-adjusted associations between snoring and incident dementia. IR, incident rate; HR, hazard ratio; CI, confidence interval. Model 1 included age, sex, ethnicity, education, and Townsend deprivation index quintiles. Model 2 was further adjusted for smoking status, alcohol consumption, body mass index, daytime dozing, depression, diabetes, hypertension, and cardiovascular diseases.

Snoring was more strongly related to dementia in older participants (Figure 3). The HRs for dementia were 0.87 (95% CI 0.82 to 0.92) for participants aged 65 years or older, compared to 0.98 (95% CI 0.91 to 1.06) for those under 65 years old (*p*_interaction_<0.001). Larger magnitude of association was also observed for individuals who were *APOE* ε4 carriers compared to non-carriers (0.93 [95% CI 0.87 to1.00] vs 0.94 [0.88 to 1.01]; *p*_interaction_=0.004). No significant interaction effect was observed between snoring and sex (*p*_interaction_>0.05).

**Figure 3.**
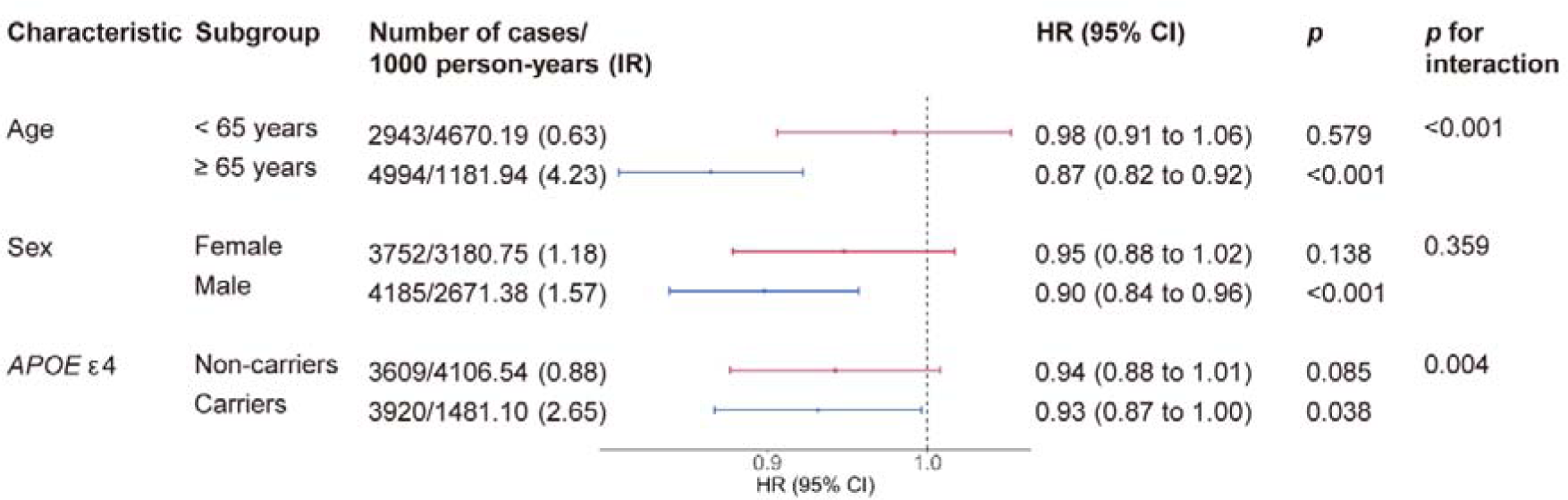
Subgroup analyses of the association between snoring and incident all-cause dementia. IR, incident rate; HR, hazard ratio; CI, confidence interval. The models were adjusted for age, sex, ethnicity, education, Townsend deprivation index quintiles, smoking status, alcohol consumption, body mass index, daytime dozing, depression, diabetes, hypertension, and cardiovascular diseases.

The association between snoring and risk of dementia decreased with increasing length of follow-up, with HRs of 0.79 (95% CIs 0.67 to 0.94), 0.89 (95% CIs 0.82 to 0.96), and 0.96 (95% CIs 0.90 to 1.03) for a follow-up length of ≤5 years, 5 to 10 years, and over 10 years, respectively (e-Figure 1). The associations between snoring and dementia remained largely unchanged after excluding participants with a history of sleep apnoea at baseline or imputing missing exposure and covariate data (e-Figure 2 and e-Table 4).

### MR analysis

In the forward direction, 31 SNPs were selected as genetic instruments for snoring (mean *F-statistic* is 40.9), with no outliers detected using Radial-MR (e-Figure 3). Genetically predicted snoring was not causally associated with the risk of AD in the IVW and all sensitivity analyses (Table 2). In the reverse direction, 30 SNPs were selected as genetic instruments for AD (mean *F-statistic* is 105.2), with no outliers detected using Radial-MR (e-Figure 4). While the IVW analysis was non-significant, diagnostic tests indicated that there was evidence of both heterogeneity and pleiotropy, suggesting that the IVW estimates may be biased. Sensitivity analyses, including MR-Egger (odds ratio [OR], 0.994; 95% CI 0.990 to 0.998), p = 0.004), WME (0.995, 95% CI 0.991 to 0.999, p = 0.013), and WMBE (0.996, 95% CI 0.992 to 0.999, p = 0.031), indicated that increased genetically predicted AD was causally associated with a reduced risk of snoring.

**Table 2.** Bi-directional causal estimates between snoring and Alzheimer’s disease.

|  | **Snoring → AD (Univariate MR)** |  | **AD → Snoring (Univariate MR)** |  | **AD → Snoring (Multivariable MR)** |  |
| --- | --- | --- | --- | --- | --- | --- |
| **Method** | **OR (95% CI)** | ***p*** | **OR (95% CI)** | ***p*** | **OR (95% CI)** | ***p*** |
| IVW | 0.838 (0.387 to 1.817) | 0.655 | 0.998 (0.995 to 1.001) | 0.114 | 1.002 (0.994 to 1.011) | 0.616 |
| MR-Egger | 23.829 (0.255 to 2223.326) | 0.181 | 0.994 (0.990 to 0.998) | 0.004 | 1.004 (0.995 to 1.013) | 0.362 |
| WME | 1.144 (0.357 to 3.667) | 0.821 | 0.995 (0.991 to 0.999) | 0.013 | 1.000 (0.993 to 1.008) | 0.912 |
| WMBE | 1.230 (0.147 to 10.268) | 0.850 | 0.996 (0.992 to 0.999) | 0.031 | 1.009 (0.997 to 1.021) | 0.132 |
| **Diagnostics** | **Estimate** | ***p*** | **Estimate** | ***p*** | **Estimate** | ***p*** |
| F-statistic | 40.9 | NA | 105.2 | NA | 13.2 | NA |
| Cochran's Q | 40.6 | 0.095 | 32.9 | 0.282 | 252.1 | $6.04 \times 10^{-19}$ |
| MR-Egger Intercept | -0.025 | 0.151 | 0.001 | 0.009 | 0.001 | 0.031 |

AD, Alzheimer's disease; MR, Mendelian randomization; OR, odds ratio; CI, confidence interval; IVW, inverse variance weighted; WME, Weighted Median Estimator; WMBE, Weighted Mode Based Estimator; NA, not applicable.

For MVMR, a total of 86 SNPs were selected as genetic instruments for BMI (N_SNP_ = 67) and AD (N_SNP_ = 19), with conditional *F-statistics* of 45.4 and 13.2 respectively. In the MVMR-IVW analyses, the causal association between genetically predicted AD and snoring was attenuated (Table 2). There was evidence of heterogeneity, and sensitivity analyses indicated that genetically predicted BMI remained significantly associated with an increased risk of snoring (e-Figure 5).

## Discussion

In this study, we investigated the longitudinal association between snoring and dementia in 450,027 participants from UK Biobank. Our results showed that snoring is associated with a lower risk of both all-cause dementia and AD, while no significant association was observed with VaD. Furthermore, this association was stronger in older individuals, *APOE* ε4 allele carriers, and during shorter follow-up periods. We further explored the relationship between snoring and AD using univariate MR analyses, which revealed a potential causal effect of AD on the risk of snoring but no significant evidence for a causal effect of snoring on AD. Our multivariable MR analyses suggested that the association between AD and snoring was mainly driven by BMI. The results indicate that the phenotypic association between snoring and decreased dementia risk is likely the result of reverse causation, possibly mediated by decreased BMI levels during the preclinical phase of dementia.

Our findings suggest that the association between snoring and all-cause dementia and AD may be attributed to reverse causation. In our study, the median follow-up time was 13.5 years; however, the accumulation of AD pathology in the brain can occur more than 20 years before clinical symptoms onset.^22^ Therefore, although we excluded participants with clinically diagnosed dementia at baseline, it is plausible that many remaining individuals are in the prodromal phase of AD, particularly the older individuals and those carrying the *APOE* ε4 allele, in whom we observed a stronger negative association between snoring and dementia. This complements previous studies that found an association between snoring and higher cognitive function among older adults over a shorter follow-up period of 10 years.^9^ In contrast, no such association was found in a relatively younger sample (mean 52.3 years) with a longer follow-up of 22.5 years.^23^ A previous cross-sectional study of non-demented older adults also found that *APOE* ε4 carriers reported less snoring than non-carriers.^24^ The direction of the phenotypic association between snoring and dementia was further supported by our univariate MR results. Specifically, we identified a one-way causal relationship between AD and a reduced risk of snoring, which survived in multiple sensitivity analyses.

The role of BMI, one of the strongest predictors of snoring,^1^ is of significant interest in elucidating the relationship between AD and snoring. Indeed, our multivariable MR results suggested that the association between AD and snoring was driven by BMI. Specifically, a lower BMI, as frequently observed during the preclinical phase of AD,^16, 25^ may lead to a reduced risk of snoring in prodromal AD. Studies have established a dynamic association between BMI and dementia across the life course, known as the “obesity paradox”. Obesity in midlife, especially before the age of 50, has been linked to an increased risk of dementia, whereas in late-life, the association between high BMI and dementia often reverses.^26, 27^ Prior MR studies have revealed that genetic liability to AD is linked to lower BMI,^28, 29^ highlighting the role of late-life weight loss as a prodromal factor for AD. Accumulating evidence suggests that AD pathology leads to impaired functions of the hypothalamic and other brain regions crucial for metabolic regulations, which contribute to weight loss in the early stages of AD.^30, 31^ Future research should explore how BMI or change in BMI at different stages of life may influence the association between snoring and risk of dementia.

Notably, snoring is commonly considered as a sign of obstructive sleep apnoea (OSA), which has been linked to an increased risk of cognitive impairment.^12, 13^ Contrary to these findings, our current study suggested an association between snoring and a decreased risk of dementia. It is important to note that 60%-80% of snorers do not exhibit apnoea severe enough to warrant an OSA diagnosis, a condition often referred to as “simple snoring”.^1^ To account for the potential impact of OSA on the relationship between snoring and dementia, we performed a sensitivity analysis by excluding participants with a history of sleep apnoea at baseline, and the results remained unchanged. Our results suggest that snoring and OSA may have distinct clinical implications for dementia.

In our MR study, we found no causal association between snoring and risk of AD. It is important to note that snoring, through hypoxia and inspiratory vibrations, may contribute to vascular pathology and thus have a more significant impact on the risk of VaD.^32, 33^ Meanwhile, there are no well-established mechanisms to explain a potential link between snoring and risk of AD. Our observational study did not identify an association between snoring and VaD. This lack of association may be attributed to the diagnostic criteria for VaD, which have low sensitivities compared to AD,^34^ leading to misclassification that biased estimates towards the null. The limited sample size of VaD cases in most studies also constrained statistical power and the ability to achieve statistical significance.

This study has several limitations. Firstly, self-reported snoring may be subject to recall bias, which could lead to misclassification of snorers and non-snorers and potentially bias the results. Secondly, dementia cases were identified using UK Biobank linked hospital inpatient records and death registry. This approach may result in the underestimation of milder forms of dementia. Furthermore, the utilization of these two sources yielded low positive predictive values for VaD (33.3%), compared with all-cause dementia (84.5%) and AD (70.8%).^35^ The potential misclassification of dementia, particularly in the case of VaD, should not be disregarded. Thirdly, the lack of a published GWAS of VaD limited our ability to examine the causal relationship between snoring and VaD.

In our study, we found that snoring, a common condition in older adults, was associated with a lower risk of all-cause dementia and AD, particularly in older individuals and *APOE* ε4 carriers. Our MR analyses suggest potential reverse causation, where genetic liability to AD was associated with reduced snoring, possibly through lower BMI in prodromal AD. Future studies are needed to clarify the causal relationship between snoring and different subtypes of dementia and to elucidate underlying mechanisms. The role of BMI should be carefully considered in research on AD and snoring.

## Declarations

### Ethics approval

The North West Multi-centre Research Ethics Committee granted approval to UK Biobank, and UK Biobank obtained informed consent from all participants.

## Data Availability

The UK Biobank data is accessible online at https://www.ukbiobank.ac.uk for researchers who have received approval for their proposals of data use from the UK Biobank.

## Supplementary data

Supplementary data are available at IJE online

## Author Contributions

Conception and design of the study: Y.L., S.A., Y.G. Acquisition and analysis of data: Y.G., S.A., Y.L. Manuscript drafting: Y.G., S.A., Y.L. Manuscript editing: Y.G., S.A., Y.L, W.B., C.A.R., K.Y.

## Funding

Yaqing Gao is funded through Jardine-Oxford Graduate Scholarship. Dr. Yue Leng is supported by the National Institute on Aging (NIA) (R00AG056598).

## Competing Interests

None declared.

## Notes

### Competing Interest Statement

The authors have declared no competing interest.

### Author Declarations

The North West Multi-centre Research Ethics Committee granted approval to UK Biobank.

