## Supplementary materials for "Snoring and risk of dementia: a prospective cohort and Mendelian randomization study"

**Supplement**

### e-Table 1. International Classification of Disease codes in the hospital inpatient records and death registries

| **Disease** | **ICD-9 codes** | **ICD-10 codes** |
| --- | --- | --- |
| All-cause dementia | 2902, 2903, 2904, 2912, 2941, 3310, 3311, 3312, 3315 | A810, F00, F01, F02, F03, F051, F106, G30, G310, G311, G318, I673 |
| Alzheimer’s disease | 3310 | F00, G30 |
| Vascular dementia | 2904 | F01, I673 |
| Sleep apnoea | 3272 | G473 |
| Depression | 2962, 2963, 29682 | F32, F33 |
| Diabetes (excluding gestational diabetes) | 250 | E10-E11 |
| Hypertension | 401-405 | I10-I15, H35.03, I67.4 |
| Cardiovascular disease | 410-415, 425, 4273, 428, 430, 431, 434, 436 | I20-I26, I42, I43, I48, I50, I60, I61, I63, I64 |

### e-Table 2. Definition of variables included in the UK Biobank data showcase

| **Variable** | **UK Biobank field ID(s)** | **Measurement item(s)** | **Definition/categorization** |
| --- | --- | --- | --- |
| Age | 34, 53 | (Registry, updated by participant) year of birth and assessment | Age = year of assessment – year of birth |
| Sex | 31 | (Registry, updated by participant) |  |
| Ethnicity | 21000 | (Questionnaire) amalgam of sequential branching questions | We used the first non-missing value in all measurement occasions.   1) White; 2) Non-white (Asian or Asian British, Black or Black British, Chinese, Mixed, and other ethnic group) |
| Education | 6138 | (Questionnaire) self-reported qualifications | Participants who obtained multiple qualifications were assigned the highest qualification achieved, which was converted to the International Standard Classification for Education (ISCED) coding for years of education ^1,2^:   1) Tertiary: College/University Degree, National Vocational Qualification (NVQ) / Higher National Diploma (HND) / Higher National Certificate (HNC) or equivalent 2) Post-secondary non-tertiary: Other professional qualifications (e.g., nursing, teaching) 3) Secondary: General Certificate of Education Advanced Level (A levels) / General Certificate of Education Advanced Subsidiary Level (AS levels) or equivalent, General Certificate of Education Ordinary Level (O levels) / General Certificate of Secondary Education (GCSEs) or equivalent, Certificate of Secondary Education (CSEs) or equivalent 4) Primary: None of the above |
| Townsend deprivation index (TDI) | 189 | TDI was calculated immediately prior to participant joining UK Biobank. Each participant is assigned a score corresponding to the preceding national census output area in which their postcode is located. | The whole UK Biobank sample was divided into quintiles based on their TDI, arranged from richest (quintile 1) to poorest (quintile 5). |
| Smoking status | 20116 | (Questionnaire) self-reported smoking-status | 1) Never; 2) Previous; 3) Current |
| Alcohol consumption | 20117, 1558 | (Questionnaire) self-reported drinking status and drinking frequency | 1) ≤ 4 times week; 2) Daily or almost daily ^3^ |
| Daytime dozing | 1220 | (Questionnaire) self-reported sleep-related traits | 1) Never/rarely; 2) Sometimes/often/all the day |
| Body mass index (BMI) | 21001 | (Physical measures) height and weight | The classification for BMI is in use by the World Health Organization ^4^:   1) Normal (<25 kg/m^2^); 2) Overweight (25–30 kg/m^2^); 3) Obese (≥30 kg/m^2^) |
| History of dementia | 20002 | (Interview) self-reported illness | "dementia/alzheimers/cognitive impairment" |
| History of sleep apnoea | 20002 | (Interview) self-reported illness | "sleep apnoea" |
| History of depression | 20002 | (Interview) self-reported illness | "depression" |
| History of diabetes (excluding gestational diabetes) | 20002 | (Interview) self-reported illness | "diabetes", "type 1 diabetes", "type 2 diabetes" |
| History of hypertension | 20002 | (Interview) self-reported illness | "hypertension", "essential hypertension" |
| History of cardiovascular disease | 20002 | (Interview) self-reported illness | "angina", "heart attack/myocardial infarction", "cardiomyopathy", "hypertrophic cardiomyopathy (hcm / hocm)", "heart failure/pulmonary oedema", "atrial fibrillation", "brain haemorrhage", "ischaemic stroke", "stroke", "subarachnoid haemorrhage", "transient ischaemic attack (tia)", "pulmonary embolism +/- dvt", "deep venous thrombosis (dvt)" |

### e-Appendix 1. Genotyping, multiple imputation, and Mendelian Randomization

*APOE* genotypes were estimated from 2 single nucleotide polymorphisms (SNP), rs7412 and rs429358. Participants with at least one copy of *APOE* ε4 were considered *APOE* ε4 carriers. Ambiguous genotypes, *APOE* ε1/ ε3 and *APOE* ε2/ε4, were excluded.

We used multiple imputation by chained equations to impute missing neuroticism score and covariates. The imputation model included the exposure (self-reported snoring), all preselected covariates, the Nelson-Aalen estimate of cumulative baseline hazard, and dementia status.^5^ We conducted Cox proportional hazards regression analyses to assess the association between snoring and incident dementia and its subtypes in five imputed datasets, and the results from each of the five imputed datasets were combined using Rubin’s rules.

To identify instruments for each phenotype, genome-wide significant SNPs were extracted (p < 5 × 10^−8^) from their respective GWASs and clumped (r^2^ < 0.001, 10mb window, EUR reference population). SNPs for each exposure were subsequently extracted from each outcome GWAS, with SNPs that were unavailable in the outcome GWAS summary statistics replaced with proxy SNPs that were in high genetic linkage (r^2^ ≥ 0.8, 1000 Genomes European reference population). From the full list of SNPs associated with both AD and BMI, we first obtained proxy variants for variants not present in each respective exposure and then performed a second clumping procedure to obtain independent variants (r^2^ < 0.001 within 10mb distance). The exposure–outcome datasets were harmonized to ensure that SNP effect for the exposure and outcome were relative to the same allele, allowing forward-strand ambiguous SNPs to be inferred using allele frequency information, with strand-ambiguous SNPs with intermediate allele frequencies (AF > 0.42) removed from the analysis. Variants in the APOE region (CHR:BP 19:44912079-45912079) were excluded.

Code for implementing the Mendelian randomization analyses is available at: <https://github.com/AndrewsLabUCSF/Snoring-AD-MR>. All statistical analyses were conducted in R (v4.2.2) using TwoSampleMR (v0.5.6), MVMR (v0.3), RMVMR (v0.2), and MendelianRandomization (v0.7.0).

### e-Table3. Harmonized datasets for two-Sample MR analysis of the association between snoring and AD

See e-Table3.csv

### e-Figure 1. Association between snoring and incident all-cause dementia by different follow-up periods

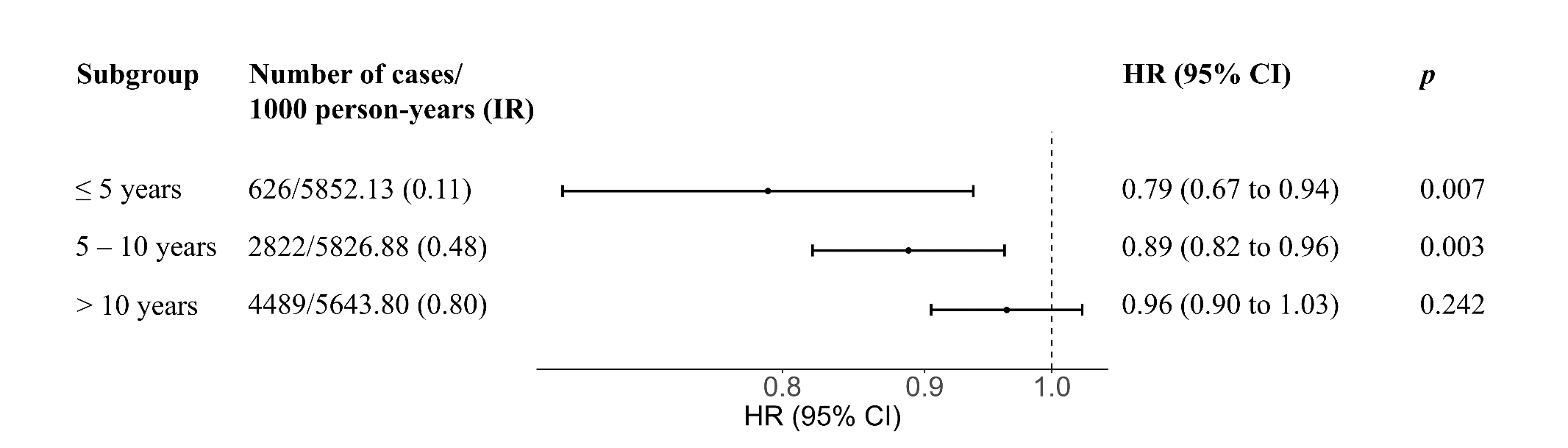

IR, incident rate; HR, hazard ratio; CI, confidence interval. The models were adjusted for age, sex, ethnicity, education, Townsend deprivation index quintiles, smoking status, alcohol consumption, body mass index, daytime dozing, depression, diabetes, hypertension, and cardiovascular diseases.

### e-Figure 2. Association between snoring and incident dementia after excluding participants with a history of sleep apnoea at baseline

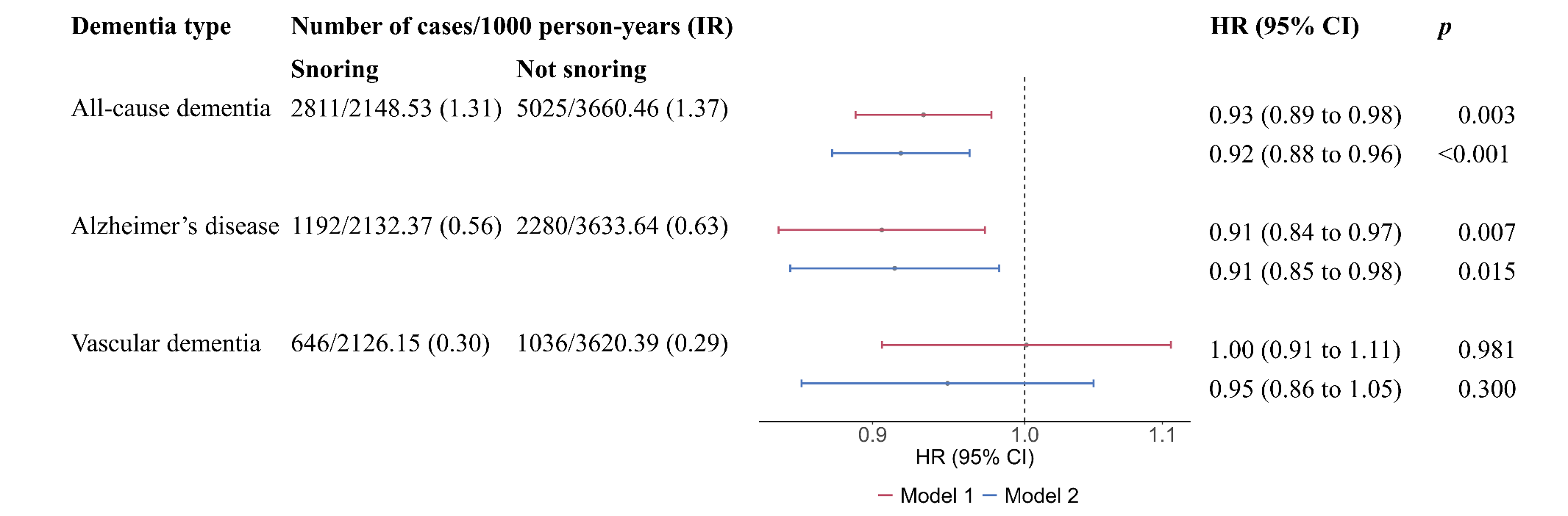

IR, incident rate; HR, hazard ratio; CI, confidence interval. Model 1 included age, sex, ethnicity, education, and Townsend deprivation index quintiles. Model 2 was further adjusted for smoking status, and alcohol consumption, body mass index, daytime dozing, depression, diabetes, hypertension, and cardiovascular diseases.

### e-Table 4. Association between snoring and incident dementia after multiple imputation of missing exposure and covariate data

| **Dementia type** | **HR (95% CI)** | ***p*** |
| --- | --- | --- |
| All-cause dementia | 0.92 (0.88 to 0.96) | <0.001 |
| Alzheimer’s disease | 0.92 (0.86 to 0.98) | 0.015 |
| Vascular dementia | 0.93 (0.85 to 1.03) | 0.170 |

HR, hazard ratio; CI, confidence interval. Models included age, sex, ethnicity, education, Townsend deprivation index quintiles, smoking status, alcohol consumption, daytime dozing, depression, diabetes, hypertension, cardiovascular diseases, and BMI.

### e-Figure 3. Scatter plot (A), funnel plot (B), Radial-MR (C), and coefficient plot (D) for the univariate MR results (Snoring → AD)

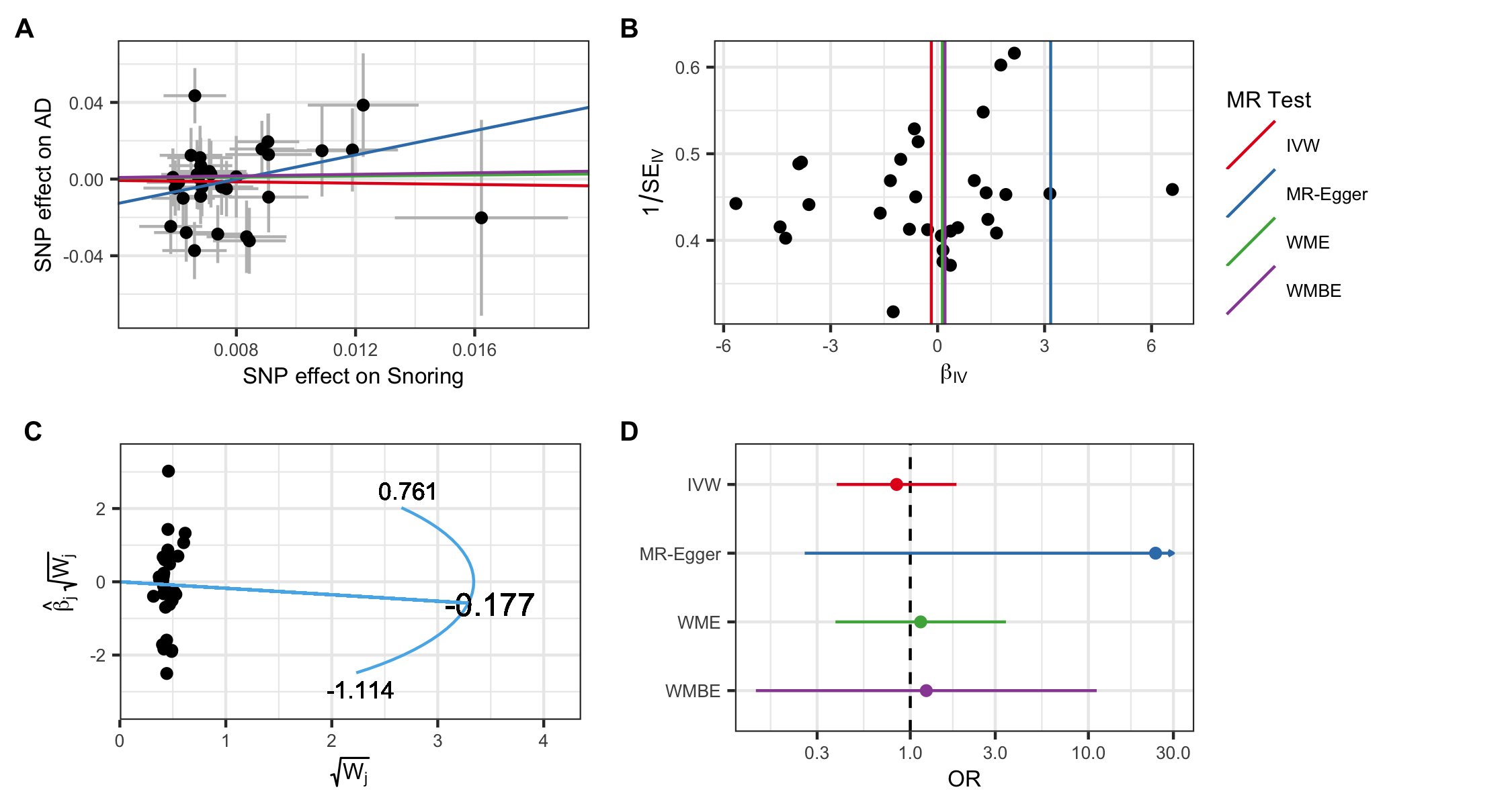

SNPs, single-nucleotide polymorphisms; IVW, inverse variance weighted; WME, Weighted Median Estimator, WMBE, Weighted Mode Based Estimator; OR, odds ratio.

### e-Figure 4. Scatter plot (A), funnel plot (B), Radial-MR (C), and coefficient plot (D) for the univariate MR results (AD → Snoring)

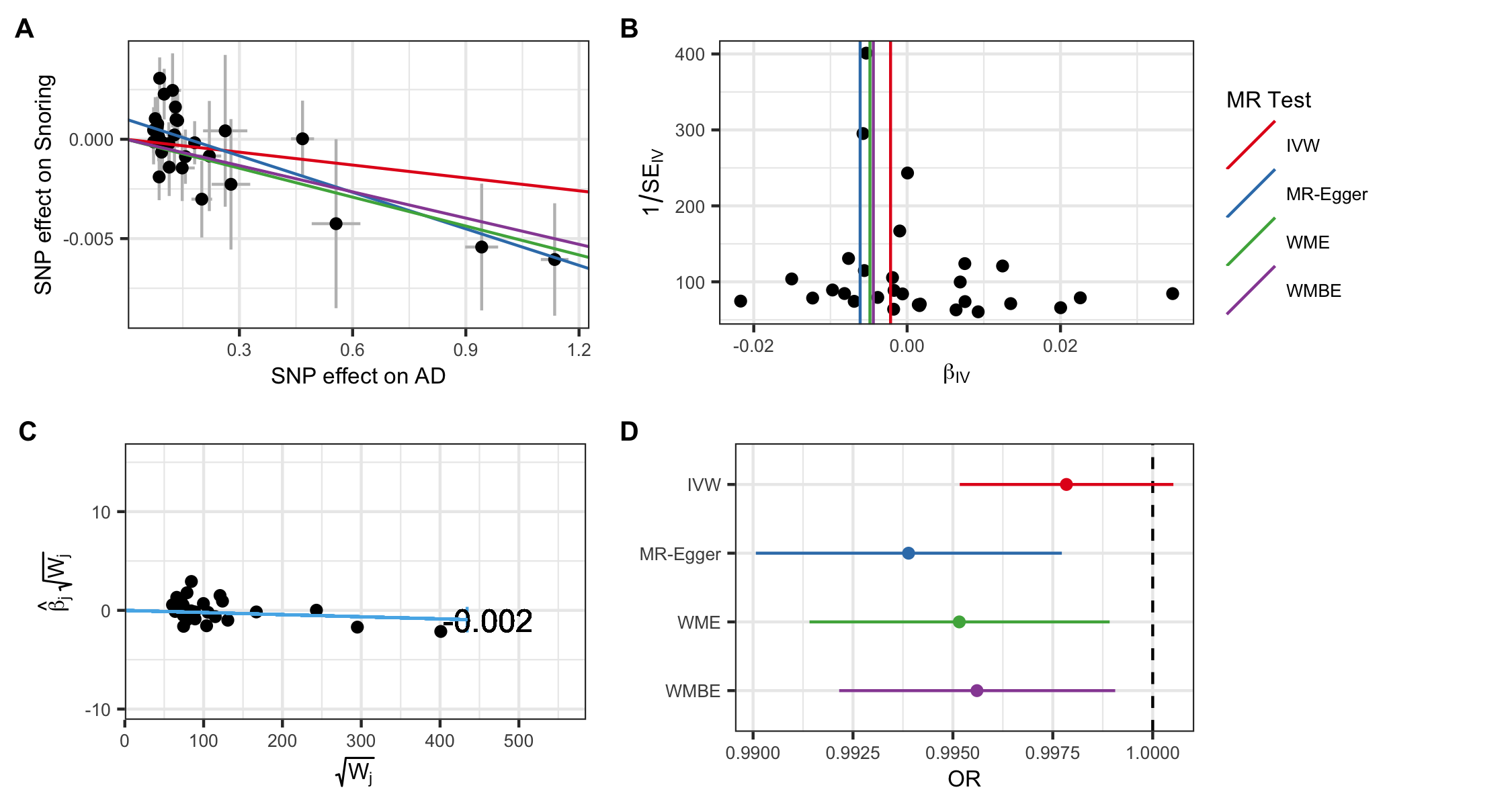

SNPs, single-nucleotide polymorphisms; IVW, inverse variance weighted; WME, Weighted Median Estimator, WMBE, Weighted Mode Based Estimator; OR, odds ratio.

### e-Figure 5. Scatter plot (A), funnel plot (B), Radial-MR (C), and coefficient plot (D) for the univariate MR results (BMI → Snoring)

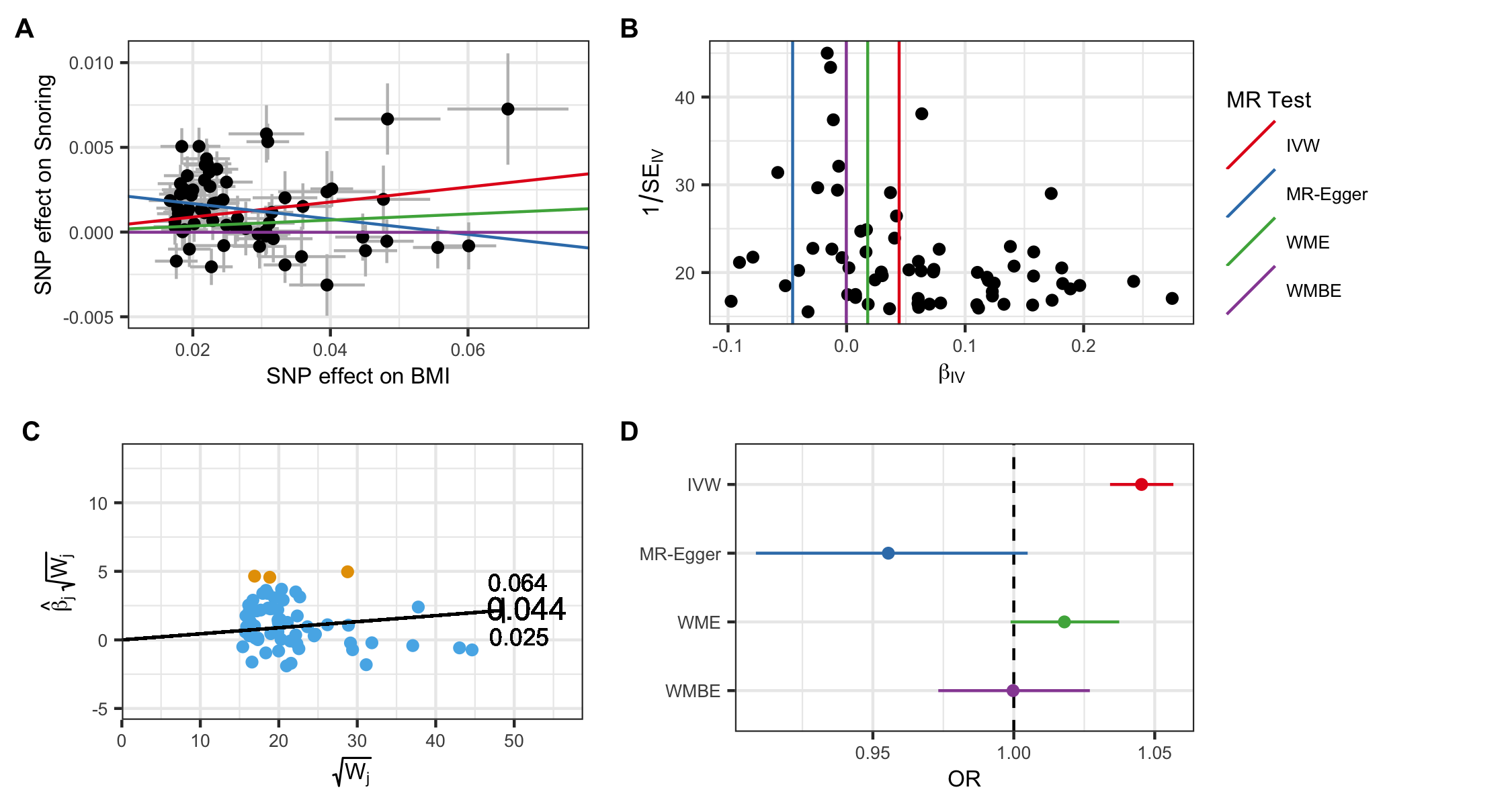

SNPs, single-nucleotide polymorphisms; IVW, inverse variance weighted; WME, Weighted Median Estimator, WMBE, Weighted Mode Based Estimator; OR, odds ratio.
